# Early-life exposure to air pollution alters resting-state functional connectivity patterns in late childhood

**DOI:** 10.1101/2025.11.16.25340328

**Authors:** Snežana Todorović, Mikolaj Compa, Daniel S. Margulies, Emilia Rutkowska, Yarema Mysak, Malgorzata Lipowska, Katarzyna Sitnik-Warchulska, Bernadetta Izydorczyk, Katarzyna Kaczmarek-Majer, Krzysztof Skotak, Anna Degórska, Clemens Baumbach, James Grellier, Iana Markevych, Marcin Szwed

## Abstract

**Background:** Adverse environmental exposures such as air pollution can disrupt brain development, contributing to long-term risk of neurodevelopmental and psychiatric disorders. Resting-state functional connectivity (rsFC) captures large-scale brain alterations seen in such disorders, and growing evidence links air pollution exposure to adverse cognitive and mental health outcomes.

**Methods:** We used connectome-based predictive modeling (CPM), a data-driven approach that uses brain states to predict individual variables, in this case, air pollution exposures, to investigate whether individual patterns of rsFC at 10-13 years in children from the NeuroSmog study are associated with prior exposure to particulate matter (PM_10_) and nitrogen dioxide (NO_2_). The tested exposure periods comprised prenatal, from birth until the 2^nd^ birthday, 2^nd^ to 4^th^ birthday, the year prior to MRI data acquisition, and the week prior to MRI data acquisition.

**Results:** Functional connectivity at 10-13 years successfully predicted exposures to PM_10_ in two subsequent early-life periods, from birth until the age of 2, and at the age of 2-4. Predictions for other exposure periods, as well as for all tested periods of exposure to NO_2_, were not significant. Higher PM_10_ exposures in both periods were associated with a decrease in network segregation and heightened functional connectivity, in particular between the default mode, ventral attention, cerebellar, and salience networks.

**Conclusions:** Since segregation between brain networks normally decreases with age, our findings suggest that early-life exposure to PM_10_ may disrupt this developmental trajectory. Exposures during a period of increased neuroplasticity can thus exert enduring effects on functional network architecture.

## Introduction

The developing brain is particularly vulnerable to harmful environmental exposures, including air pollution (1), synthetic chemicals and psychosocial stressors (2,3). These adverse influences can have lasting consequences (4–6). Air pollution exposure, in particular, has been associated with increased psychiatric emergency visits in children (7) and increased dispensation of antipsychotics, sedatives, and sleep medications (8). Beyond its well-established impacts on respiratory and cardiovascular systems (9,10), air pollution may thus contribute to developmental and psychiatric disorders (2).

Altered patterns of large-scale functional brain organization, as measured by resting-state functional connectivity (rsFC), are a consistent feature of psychiatric disorders (4,11). These alterations can be shared across diagnostic categories (12), or be disorder-specific, as observed in autism (13), anxiety, major depressive disorder (14), schizophrenia (13), bipolar disorder (14), obsessive-compulsive disorder (14), post-traumatic stress disorder (15), and attention-deficit/hyperactivity disorder (ADHD) (13). While prior research mostly focused on the consequences of disrupted rsFC, investigating causal mechanisms of environmental origins of such alterations would inform policy debates about air quality and standards and help advance screening strategies and interventions aimed at identified at-risk individuals.

Mental disorders often emerge during childhood and adolescence (16–18). A consistent pattern of functional maturation in typically developing children includes increasing within-network specialization and selective long-range integration (19–21). The overall between-network segregation, for example, separation of the default mode network (DMN), engaged during internally oriented, self-referential thought (22) from sensory processing networks, increases with age. In parallel, networks supporting common processes, in particular the salience (SAL) and fronto-parietal (FP) network, become progressively more integrated (23).

Epidemiological studies increasingly link air pollution to child mental health (7,8) cognitive (24–28) and brain (1,5,6) outcomes. However, findings of structural brain studies are inconsistent, with results that vary across exposure windows, imaging measures, and cohorts (1,29,30). The findings of functional MRI studies, by contrast, are more consistent. Higher exposure to air pollution has been linked to less segregated sensorimotor networks, stronger coupling between task-positive and task-negative systems, and longitudinally, to persistent reductions in amygdala connectivity with attentional, somatomotor, and auditory networks, as well as lasting alterations in salience–medioparietal connectivity (31–33). Together, these findings suggest that functional network reorganization may mediate the effects of air pollution on neurodevelopment.

Here, we adopted connectome-based predictive modeling (CPM), a predictive, data-driven method developed to investigate associations between differences in the connectome and behavioral or clinical traits at the individual level (31). The predictive strength of CPM comes from its ability to identify networks of connections that reliably relate to the variable of interest within the sample, and to test how well these networks predict the variable in question in each of the participants (see Supplementary Materials). CPM has been used across diverse psychiatric and cognitive domains (32–41) to predict cognitive and clinical outcomes. Here, we applied CPM to test whether individual scores of exposure to particulate matter with aerodynamic diameter ≤10 μm (PM₁₀) and nitrogen dioxide (NO₂) could be associated with patterns of rsFC measured at ages 10-13. Specifically, we tested the the impact of early life exposure (prenatal, from birth to 2 years, and 2-4 years), a year preceding MRI data acquisition (42), as well of recent exposure, in the week prior to MRI data acquisition (24) (see Figure 1; for a previous study that addressed susceptibility windows of brain vulnerability to air pollution exposure, see (43)). Based on prior evidence that early childhood represents a critical window of heightened neurodevelopmental sensitivity to environmental exposures (44), we hypothesized that exposure during the first four years of life would show the strongest associations with functional brain organization.

**Figure 1.**
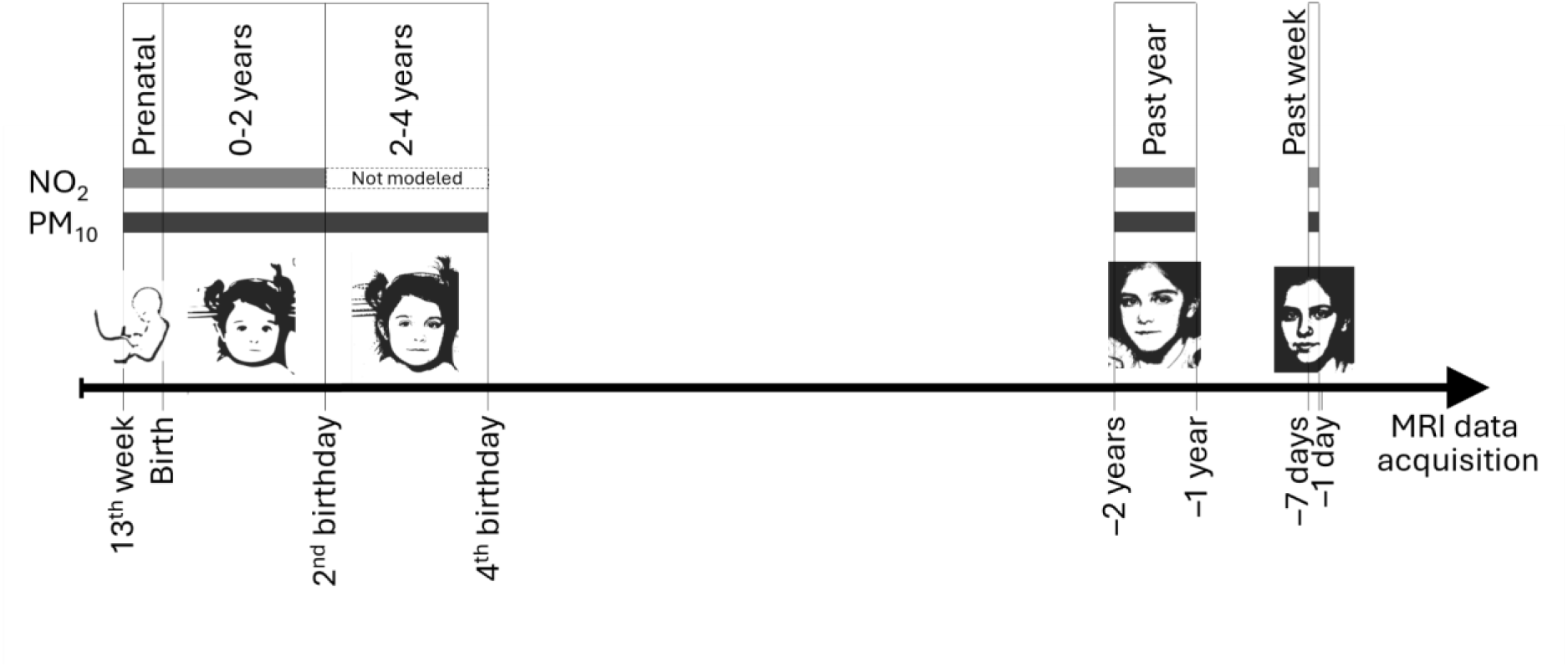
Modeled periods of exposure for particulate matter with aerodynamic diameter ≤10 μm (PM₁₀) and nitrogen dioxide (NO₂). Given the high correlations (> .9) of NO_2_ exposures over consecutive years, no model was created for NO₂ at 2-4 years. Photographs taken by the authors, modified with IA and reproduced with permission.

## Methods

The analyses were pre-registered (45). The study was approved by the ethics committee of the Institute of Psychology, Jagiellonian University, Poland (#KE_24042019A). Written informed consent was obtained from participants and their legal guardians, as detailed in the protocol article(46).

### Participants

The Neurosmog study consists of 741 children, aged 10-13 years at the time of MRI scanning, that were recruited from 18 Polish towns selected based on air pollution levels and population size (46). Briefly, typically developing children were randomly selected from local schools (stratified random sampling), while children at risk for ADHD were recruited through convenience sampling via referrals or parental applications. ADHD diagnoses were based on ICD-11 criteria and a standardized test battery, applied by field clinical psychologists to all children, and independently verified by senior clinical psychologists. Exclusion criteria included major neurological, psychiatric, or developmental conditions, low birth weight, and insufficient proficiency in the Polish language. In the present study, children with available resting state fMRI data were considered for further analysis (N = 639), of which 636 provided complete demographic data. The application of further exclusion criteria (excessive movement, missing covariate measures, and missing air pollution exposure data) resulted in a final study sample of between 348 and 370 children depending on the air pollution exposure period (see “Air pollution exposure assessment”) (Figure 2). Table 1 shows demographic data for the 370 children.

**Figure 2.**
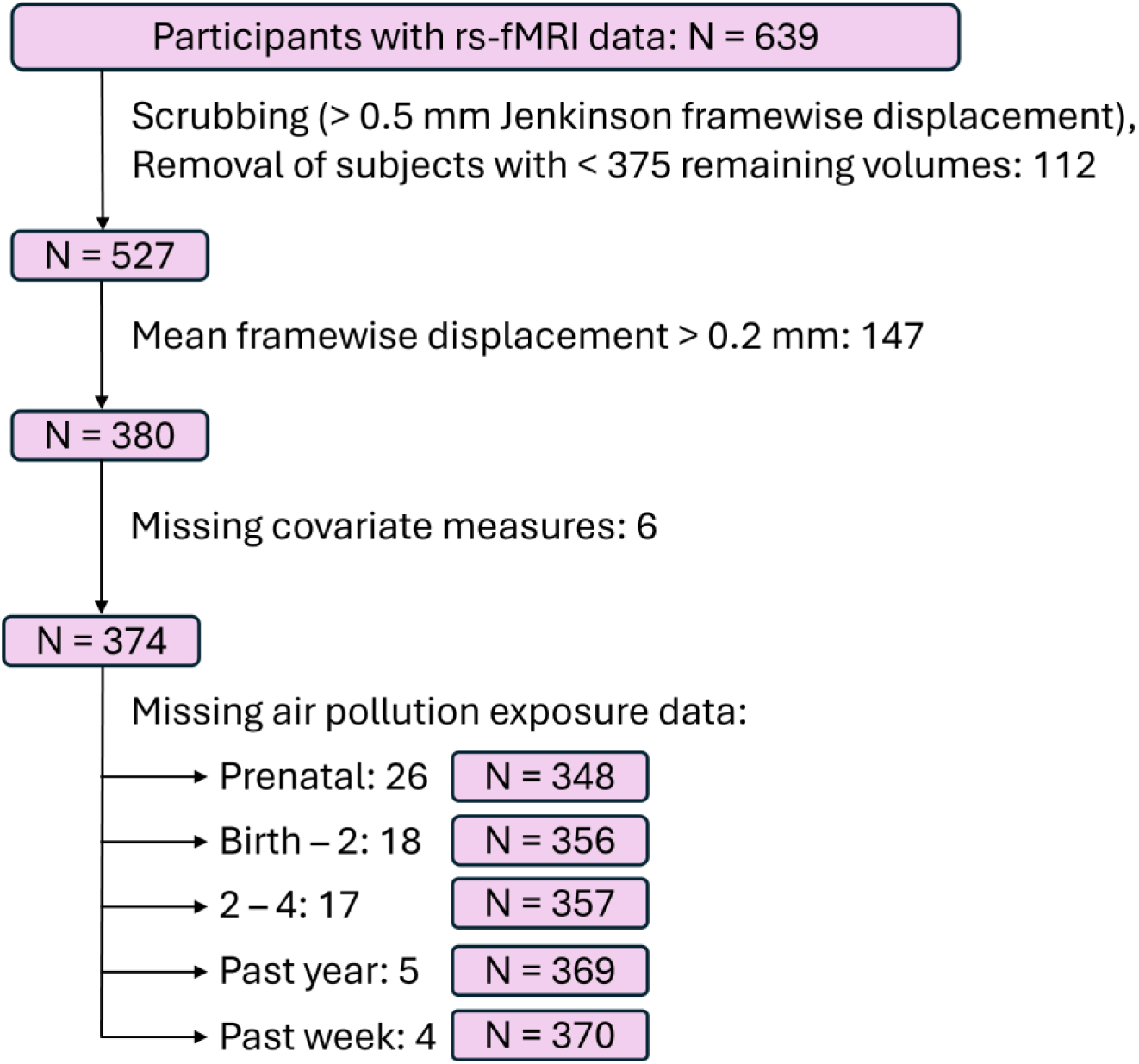
rs-fMRI data preprocessing flowchart.

**Table 1.** Demographic characteristics of the participants included in the analysis. Age – age in years at the time of MRI data acquisition, Sex – as assigned at birth, Town size – large for towns with over 90 000 inhabitants, otherwise small.

|  | Age | Sex |  | Town size |  |
| --- | --- | --- | --- | --- | --- |
|  | Mean (SD) | F | M | large | small |
| Typically Developing (N = 278) | 11.6 (0.7) | 145 | 133 | 134 | 144 |
| ADHD diagnosed (N = 92) | 11.6 (0.9) | 28 | 64 | 54 | 38 |
| Total (N = 370) | 11.6 (0.8) | 197 | 173 | 188 | 182 |

### Air pollution exposure assessment

Air pollution exposures to PM₁₀ and NO₂ were estimated at children’s home addresses using land use regression (LUR) models as detailed in prior work from our group(24,29,47). Briefly, these models integrate ground monitoring data with meteorological information, chemical transport modeling, and land-use characteristics, allowing for high-resolution, spatially refined exposure estimates suitable for epidemiological studies (48). Short-term exposures, in contrast, were derived from European Environmental Agency (EEA) monitoring data, using mean pollutant concentrations from the background station closest to each child’s residence (24). Figure 1 shows time windows for which average air pollution concentration exposures were assigned: 2^nd^ and 3^rd^ trimester of fetal development, from birth to 2^nd^ birthday, from 2^nd^ to 4^th^ birthday, for the year and for the week prior to MRI data acquisition.

### Covariates

For all exposure periods, the following covariates were included in the analysis: (1) child’s age at the time of MRI data acquisition (in months), (2) sex assigned at birth, (3) ADHD diagnosis, included to account for potential variability in functional connectivity related to clinical heterogeneity, and (4) school town type (large or small), included to account for potential differences in degree of urbanicity between smaller and larger towns. Socioeconomic status (SES) was not included as a primary covariate, as it did not correlate with air pollution exposure in our sample. Instead, SES, assessed as the minimum education level of the two parents, was examined in sensitivity analyses to assess the robustness of the observed associations. For prenatal and past-week exposure periods, an additional covariate was added, respectively birth season and time spent outdoors.

### Neuroimaging

#### Data acquisition and preprocessing

The neuroimaging data acquisition protocol is detailed elsewhere (46). In brief, the acquisition sequences for MRI and fMRI neuroimaging data were adopted from the Adolescent Brain Child Development (ABCD) study (49) and the Human Connectome Project - Development study (50). Figure 2 represents the rs-fMRI data preprocessing flowchart. Acquisition and preprocessing details are provided in Supplementary Materials.

#### Functional connectivity matrices

Single-subject whole brain rsFC matrices were calculated, using the CONN toolbox (51), between each pair of predefined functional regions of interest (ROI). 333 cortical ROI were selected from Gordon parcellation (52), 15 subcortical from FSL (53), and 26 cerebellar from the AAL (54). Thus, a total of 374 ROI was used and each participant’s functional connectivity matrix contained 374 × 374 = 139,876 values. Functional connectivity strengths were represented by Fisher-transformed bivariate correlation coefficients from a general linear model, estimated separately for each pair of ROI, characterizing the association between the time series of their blood-oxygenation-level–dependent (BOLD) signal fluctuations.

#### Connectome-based predictive modeling (CPM)

Separate CPM analyses were performed for each pollutant (PM₁₀, NO₂) and exposure window (prenatal, 0–2 years, 2–4 years, past year, and past week; see Figure 1). Because NO₂ exposure estimates were highly correlated across consecutive years (R² = 0.90–0.95, all p < .01), the 2-4-year window was excluded to avoid redundancy, resulting in nine models in total.

Each CPM followed the same cross-validation procedure. First, the sample was divided into a training set, used to identify relevant connections and estimate model coefficients, and a test set, used to evaluate model performance. Within the training set, the strength of each functional connection was correlated with individual air pollution exposure levels using two-tailed Pearson correlations, computed using all participants with available data. Connections showing a significant association (p < 0.05) were grouped into two sets: one showing positive associations, where higher exposure was linked to stronger connectivity, and one showing negative associations, where higher exposure was linked to weaker connectivity. These two sets of selected connections were stored as masks, binary matrices marking which connections were retained for each association type.

For each participant, in both training and test sets, their full connectivity matrix was reduced to a single positive and a single negative summary metric as follows. The connectivity matrix was multiplied by each mask, and the resulting connectivity values were summed and divided by two to avoid double-counting symmetric connections. This resulted in two scalar values per participant, one representing the overall strength of connections increasing with air pollution, and one for air pollution-related strength decrease. In the training set, these summary values, together with covariates, were entered into a linear regression with air pollution exposure as the outcome:

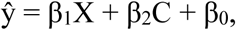

where X denotes the participant’s summary connectivity strength, C represents the covariate matrix, and ŷ is the exposure value predicted from the model. The resulting coefficients were then applied to the test set to obtain predicted exposure values. CPM significance testing, further CPM analyses methods, and exploratory analyses of the impact of air pollution exposure on functional network segregation and integration are described in Supplementary Materials.

## Results

### Predictive Performance of CPM

CPM performance metrics are shown in Table 2. Between-subject differences in functional connectivity successfully predicted PM_10_ exposure for the two early-life windows, from birth to age 2, and from age 2 to 4. In both periods, the values of PM_10_ exposure predicted by CPM significantly correlated with the values in actual data (R = 0.10, p = 0.04, and R = 0.15, p = 0.01). Predictions were based on functional connections that positively correlated with PM_10_ exposure; in other words, higher functional connectivity was predictive of higher exposures to PM_10_.

**Table 2.**
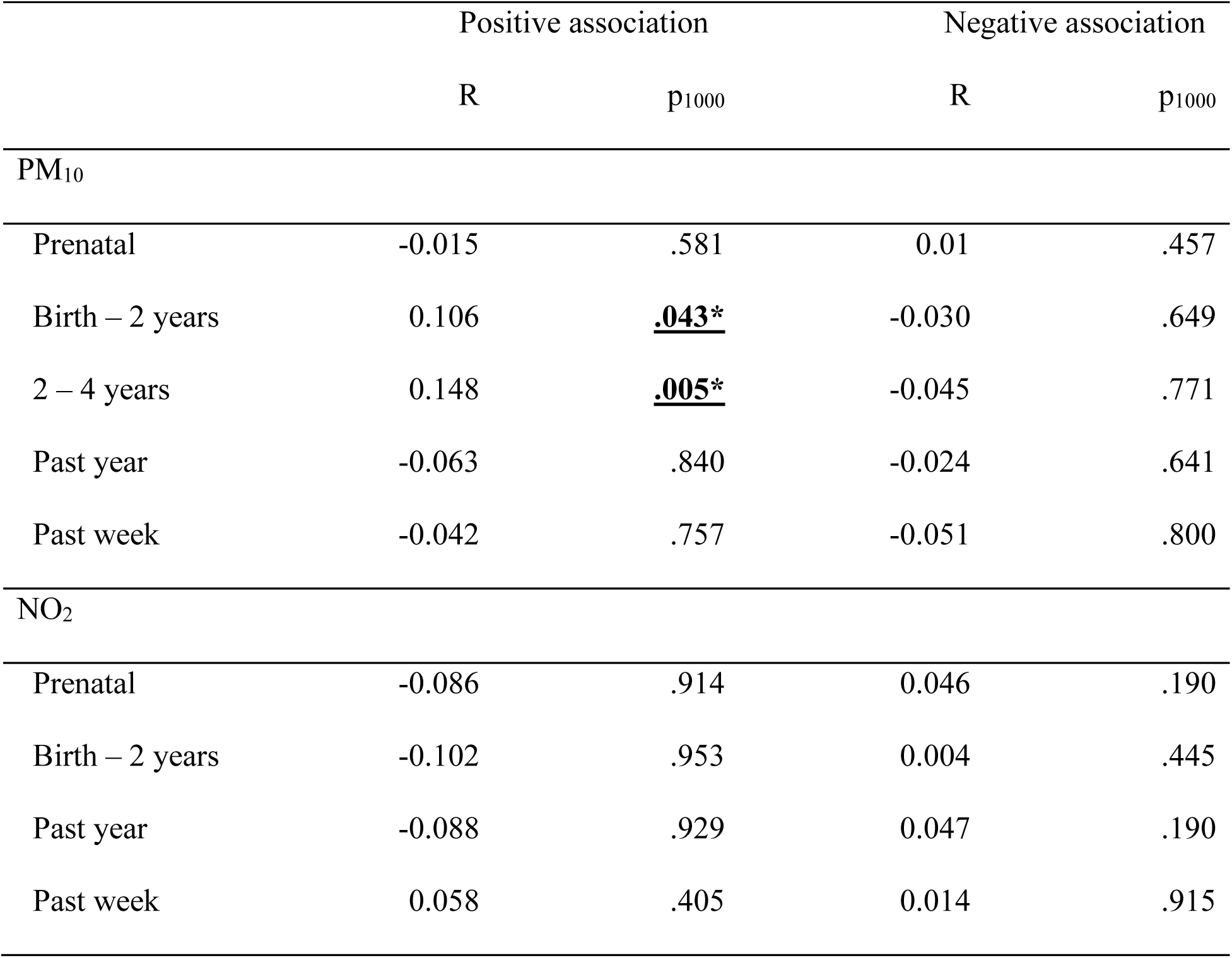
Connectome-based predictive modeling (CPM) performance. Positive and negative associations of air pollution exposure during tested time periods with functional connectivity. Positive and negative associations here signify that higher air pollution is associated with either higher or lower functional connectivity in significant (predictive) connections, respectively. R represents the correlation coefficient between the actual air pollutant exposure for the given period and the air pollutant exposure predicted by the CPM based on resting state functional connectivity. p_1000_ represents the empirical p-value for the null hypothesis R = 0 and is based on 1000 iterations of permutation testing. Statistically significant results are presented in bold.

### Distribution of predictive connections across functional networks

The CPM model identified functional connections whose strength reliably contributes to predicting individual PM_10_ exposure levels. We next examined which large-scale brain networks these predictive connections belonged to, as this reveals which systems are most affected by air pollution. Figure 3 shows predictive connections arranged by network and hemisphere. Table 3 reports the number and percentage of such predictive connections relative to all possible connections for each network. For both exposure periods (0-2 years and 2-4 years), associations were particularly evident in attentional systems, with both the dorsal and ventral attention networks (DAN, VAN) contributing alongside the default mode (DMN), cingulo-opercular (CON), cerebellar (CBL), sensorimotor (SM) and visual (VIS) networks. The networks mostly contributing to PM_10_ prediction included salience (SAL), cerebellar (CBL) and sensorimotor networks (SM), with additional contributions from VAN at 2-4 years and on the subcortical network (SUB) at 0-2 years. Figure 4 shows the distribution of these effects.

**Figure 3.**
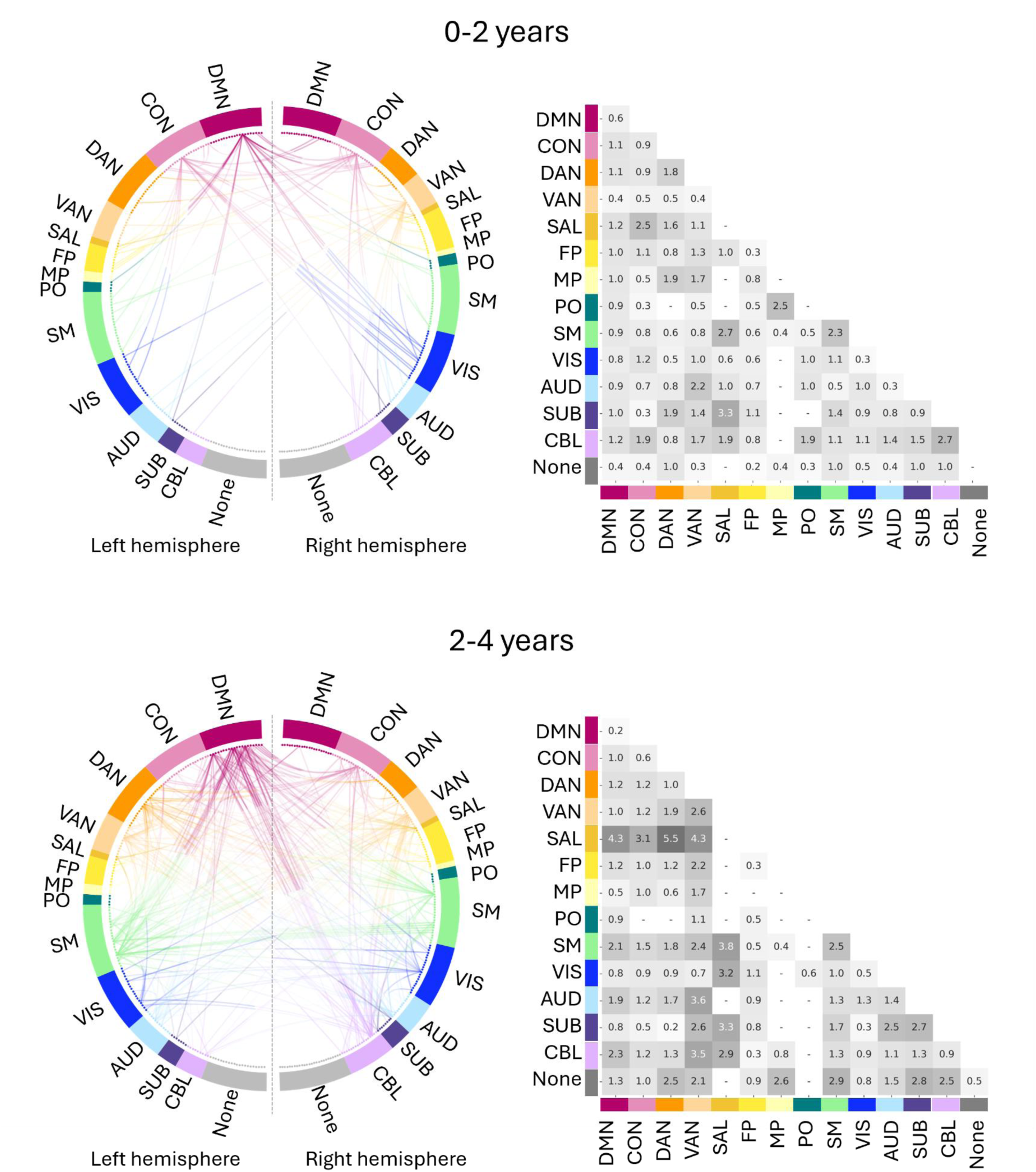
Resting state functional connections at the age 10-13 that significantly predict exposure to PM_10_ a) from birth until the age of 2 (top) and b) from the second birthday until the age of 4 (bottom). Left: Regions of interest (ROI) grouped into functional networks and arranged symmetrically by hemisphere; the length of each segment corresponds to the number ROI belonging to that network in the predefined parcellation, which can differ slightly between hemispheres. Each curve represents a functional connection between two regions that is consistently associated with PM_10_ exposure in the CPM model. To increase readability, only the functional connections of the top 25% of the most connected ROI are displayed. Right: Average percentage of connections impacted by exposure to PM_10_ per network. See Supplementary Materials for details. DMN – default, CON – cingulo-opercular, DAN - dorsal attention, VAN – ventral attention, SAL – salience, FP – fronto-parietal, MP – medio-parietal, PO – parieto-occipital, SM – sensorimotor, VIS – visual, AUD – auditory, SUB – subcortical, CBL – cerebellar, None – unassigned. PM_10_: particulate matter with a diameter of 10 μm or less, CPM: connectome-based predictive modeling.

**Figure 4.**
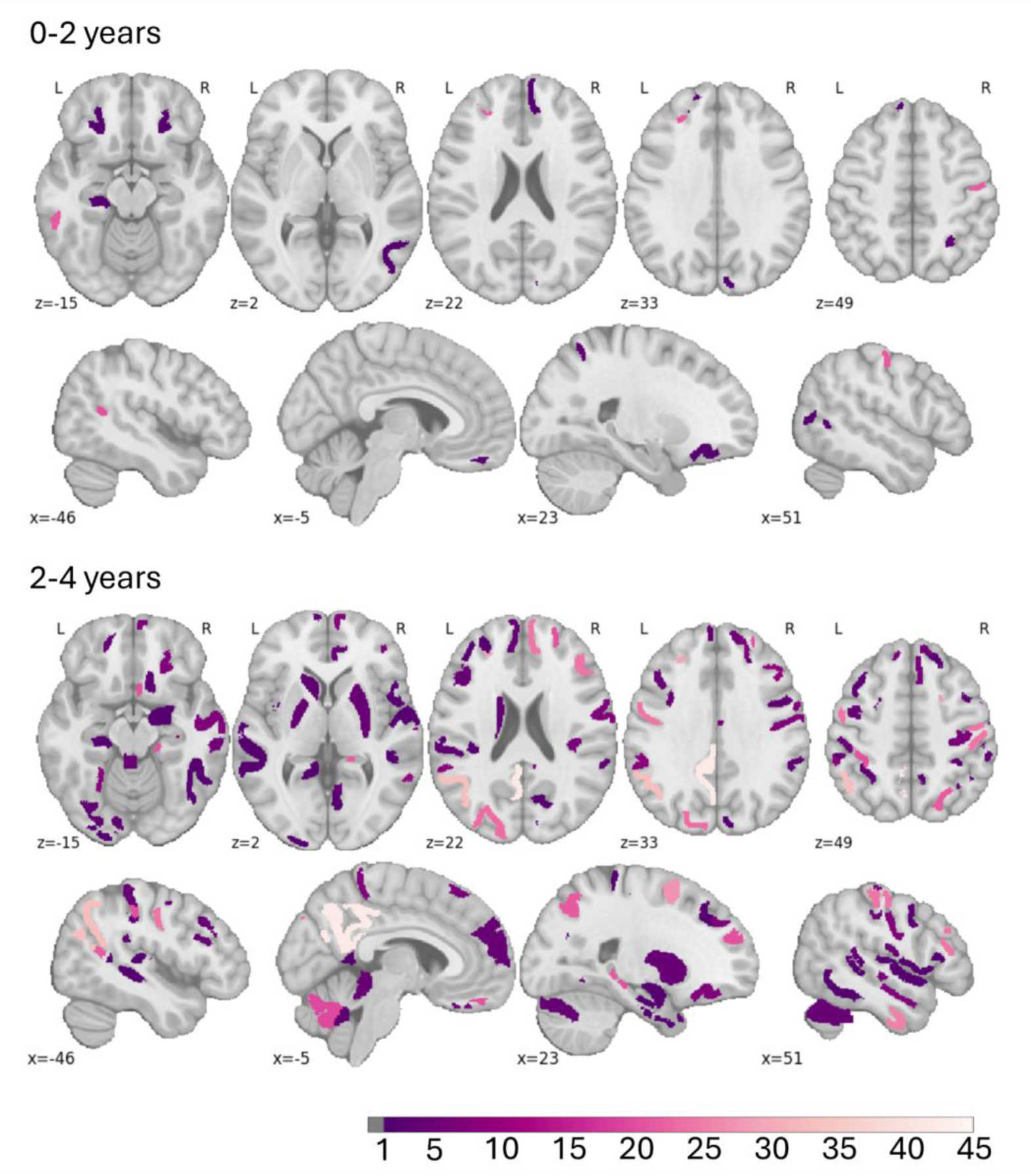
Anatomical localization of regions impacted by exposure to PM_10_ in each network a) from birth until the age of 2 (top) and b) between ages 2 and 4 (bottom). The colorbar represents each impacted regions’ degree of connectivity with other regions, i.e., the number of other regions it connects to.

**Table 3.**
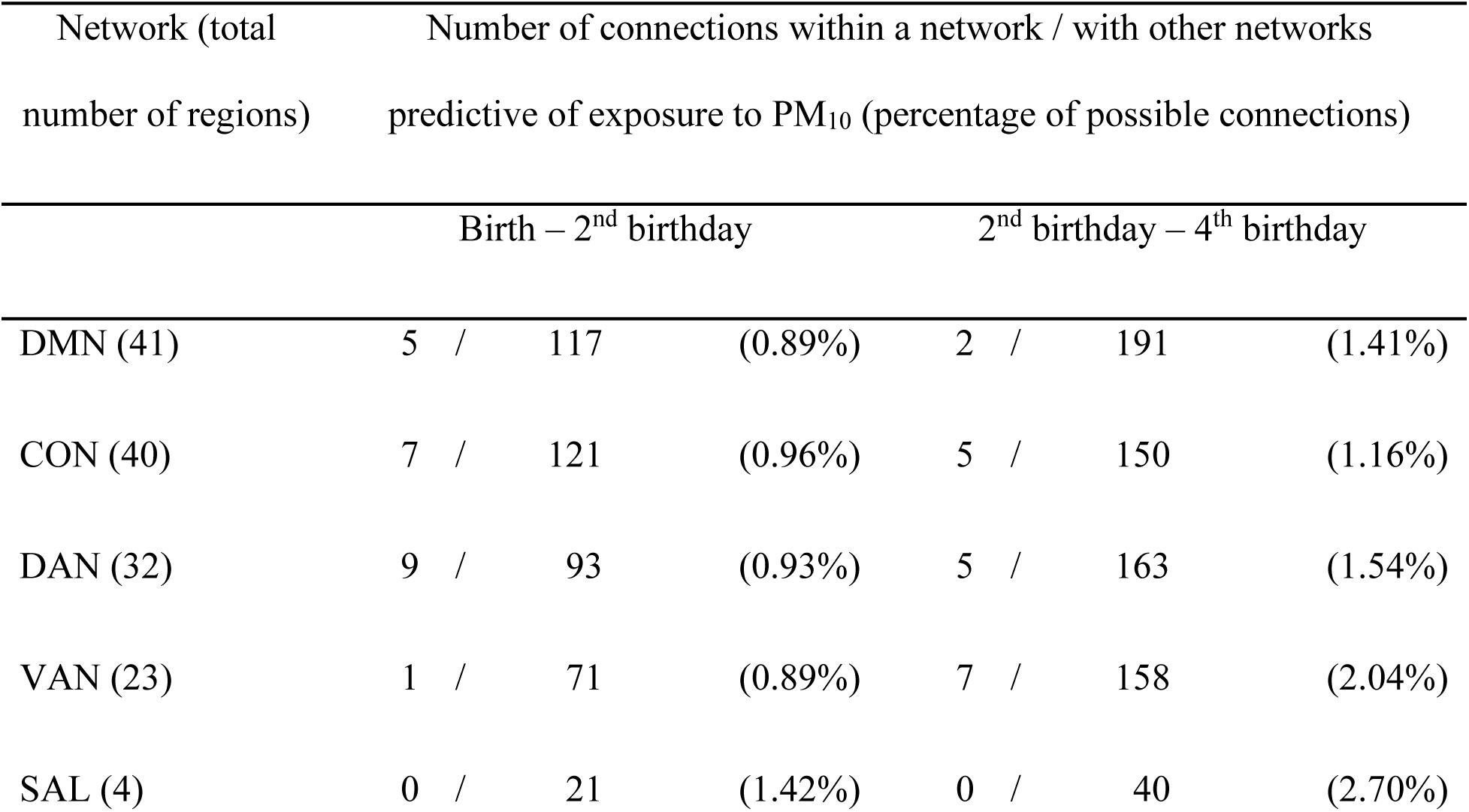
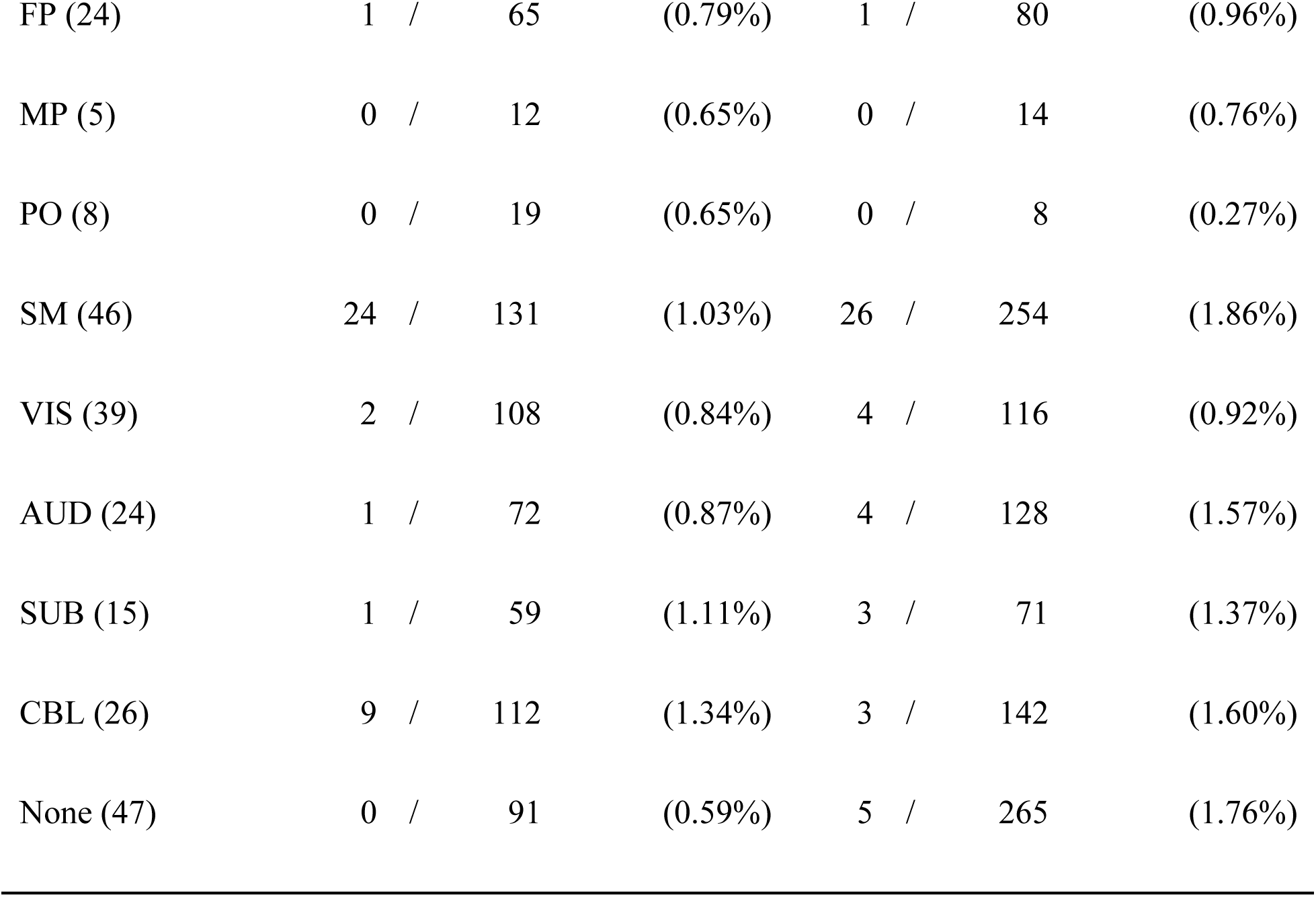
Number of connections predictive of exposure to PM_10_ per network. DMN – default, CON – cingulo-opercular, DAN – dorsal attention, VAN – ventral attention, SAL – salience, FP – fronto-parietal, MP – medio-parietal, PO – parieto-occipital, SM – sensorimotor, VIS – visual, AUD – auditory, SUB – subcortical, CBL – cerebellar, None – unassigned. PM_10_ – particulate matter with a diameter of 10 μm or less.

### Direction and strength of affected functional connections

The detected “positive” relationship between exposure to PM_10_ at 0-2 and at 2-4 years and functional connectivity values signifies that higher exposure is associated with an increase in functional connectivity values. Since for positive functional connectivity values an increase means the strengthening of the connections, and for negative functional connectivity values it means the weakening, as the negative values become closer to zero, we examined separately the positive and the negative functional connectivity values associated with increased PM_10_ exposure. As Figure 5 shows, these affected connections had predominantly positive connectivity values (median: 0.08, range: -0.31 to 0.62), suggesting weakened functional boundaries between networks and decreased segregation. While both positive and negative connections were predictive of PM_10_ exposure, positive connections had on average higher values.

**Figure 5.**
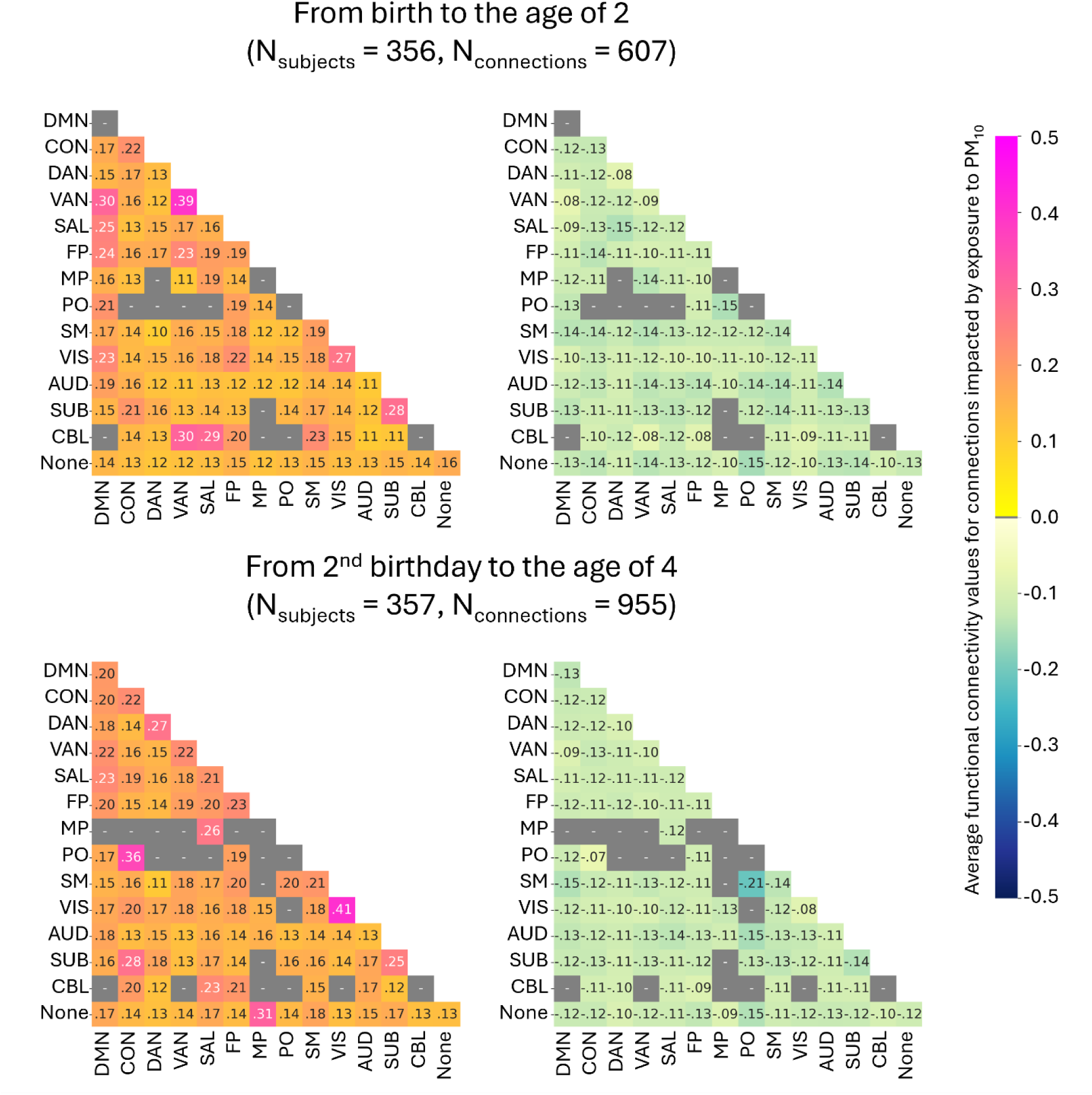
Average values of functional connections that significantly predicted PM₁₀ exposure. Left panels display subnetworks for which mean functional connectivity was positive and increased with higher PM₁₀. Right panels display subnetworks for which mean negative functional connections weakened (i.e., became less negative) with higher PM₁₀. Gray cells indicate subnetwork pairs with no suprathreshold predictive edges. The opposite association patterns (positive connectivity decreasing with PM₁₀ or negative connectivity becoming more negative) were not significant in the CPM analysis. Top row: exposure from birth until age 2; bottom row: exposure from age 2 to 4. DMN – default, CON – cingulo-opercular, DAN – dorsal attention, VAN – ventral attention, SAL – salience, FP – fronto-parietal, MP – medio-parietal, PO – parieto-occipital, SM – sensorimotor, VIS – visual, AUD – auditory, SUB – subcortical, CBL – cerebellar, None – unassigned, PM_10_ – particulate matter with a diameter of 10 μm or less, CPM – connectome-based predictive modeling.

### Network segregation

We conducted an exploratory post hoc analysis of segregation metrics for the most predictive period of PM₁₀ exposure, between ages 2 and 4 (see Supplementary Materials). Reduced segregation generally indicates less mature network organization. Higher exposure was linked to lower segregation in the default mode network (DMN), ventral attention network (VAN), and cingulo-opercular network (CON), and to a lesser extent in the parietal and visual networks (Supplementary Table 1); only DMN remained significant after correction (p < 0.001).

### Sensitivity Analysis

The results remained unchanged in a follow-up sensitivity analysis, where the calculations were repeated including SES, living in a single-parent or two-parent household, early life exposure to smoking, and maternal alcohol use during pregnancy as additional covariates. For both 0-2 years and 2-4 years periods the R values remained the same to the second decimal (R = 0.10 and 0.15) and p-values either remained the same or decreased by .01 (see Supplementary Table 2).

## Discussion

Consistent with prior reports of air pollution-related alterations in the brain development (23,42,55), we found that resting-state functional connectivity at ages 10-13 predicted individual levels of PM₁₀ exposure, with performances comparable to previous CPM applications (31) (Table 2). Our main hypothesis, that exposure between birth and age four would be most predictive, was confirmed for PM₁₀, with significant results in both the 0-2 and 2-4 year windows. Prenatal and more recent exposures were not predictive, and no associations were observed for NO₂.

Early-life exposure to PM₁₀ was mainly predicted by positive functional connectivity, which tended to increase with higher exposures (Figure 3). These predictive patterns primarily involved the default mode network (DMN), a system engaged during internally oriented, self-referential thought and typically suppressed during externally focused tasks (22); the salience (SAL) network, responsible for detecting behaviorally relevant stimuli (56); and the dorsal (DAN) and ventral attention networks (VAN) (57,58), the cingulo-opercular network (CON) (59,60), as well as the fronto-parietal network (FP) (60). In particular, higher PM₁₀ exposure increased DMN–FP coupling, both in strength (Figure 4) and in extent of affected connections (Figure 3, right), consistent with previous reports of DMN-to-FP hyperconnectivity associated with air pollution (23).

A post-hoc segregation/integration analysis suggested that increases in predicted air pollution are associated with decreased segregation of large-scale networks. This might indicate less mature network organization i.e. that higher exposure may alter the differentiation of higher-order systems developing in early childhood. This interpretation is consistent with prior work linking air pollution to less segregated attentional and sensorimotor networks (1). Our findings support the idea that early-life PM₁₀ exposure disrupts the normative trajectory of increasing network specialization.

### Developmental slowing vs. disruptive plasticity

In normative development, primary networks such as the visual system mature early, whereas higher-order systems like DAN and DMN continue to refine into adolescence (61–63), stabilizing around age 12 as networks shift from within-network cohesion toward greater between-network integration (63,64). The alterations associated with exposure to PM₁₀ we observed may reflect a range of possible developmental trajectories, including slower maturation, premature integration, or alternative reorganizations, rather than a uniform delay.

The salience network (SAL) was particularly affected in the 2-4 year exposure window, showing high proportions of predictive connections, and notably increased connectivity with DMN and FP. These networks constitute the core “triple network” in the pathophysiology of psychiatric illness. In this framework, deficits in engagement and disengagement of these networks, reflected in their altered connectivity, can account for several psychiatric disorders, in particular for psychopathologies with disturbances of cognition and the self (64).

Longitudinal studies report abuse-related within-network connectivity increases, such as neglect-related stronger SAL-DMN coupling (65–68), and similar SAL-DMN-executive shifts linked to other adversities such as unpredictability, food insecurity, and hostile environments (69–71). Comparably, PM₁₀-related increases within VAN, subcortical, and visual networks mirror findings in children with ADHD(72) and early-life psychosocial adversity (73–77), suggesting that air pollution may constitute an environmental adversity with shared neurobiological pathways, such as neuroinflammation (78,79) hypothalamic-pituitary-adrenal axis dysregulation (80), and lasting implications for cognitive control and mental health.

### Air pollution exposure as risk factor for neurodevelopmental and psychiatric conditions

Reduced segregation and atypical integration within default mode, salience, and attention networks have been reported in ADHD (81,82), ASD (83), and schizophrenia (81), and exposure to PM₁₀ and other air pollutants has been associated with increased incidence of ADHD symptoms (84,85). Prominent long-term impact of early-life PM₁₀ exposure on salience connectivity is of particular interest given the role of salience dysfunction across psychiatric conditions (86) and links between ambient air pollution and increased pediatric psychiatric emergency visits (7). Our findings support the idea that early environmental exposures may contribute to risk trajectories via alterations in core cognitive-control networks.

### Strengths and limitations

Our study has several strengths. To the best of our knowledge, it is one of the first applications of CPM to predict environmental exposures (see (87) for a recently published CPM study on exposure to PM_2.5_ in the ABCD cohort). We examined multiple early-life windows in a large, well-characterized pediatric sample. Additional strengths include high-resolution state-of-the-art assessments of air pollution exposure in a region with elevated pollution levels.

Nevertheless, several limitations should be acknowledged. The current study was cross-sectional. After splitting our sample in with and without ADHD groups, the sample sizes became too small for independent CPM analyses or between-group CPM cross-validation. Instead, we used one single subject group and included ADHD as a covariate in the CPM, which did not allow quantification of its contributions to the model.

## Conclusion

In summary, we found that early-life PM₁₀ exposure during specific developmental windows is associated with lasting differences in large-scale functional brain networks, particularly in systems supporting attention, salience, and executive control. Our results demonstrate the usefulness of predictive approaches to exposure studies, and contribute to a growing body of evidence linking environmental exposures to neurodevelopmental alterations that may increase vulnerability to psychiatric disorders.

## Data Availability

All data produced in the present study are available upon reasonable request to the authors

### Code availability

The scripts used connectome-based predictive modeling (CPM) are available at https://github.com/snezanatodorovic/CPM_CONN_MATLAB. The repository includes code and documentation sufficient to reproduce the main analyses presented in this study.

## Acknowledgments

This work was supported by “NeuroSmog: Determining the impact of air pollution on the developing brain” (Nr. POIR.04.04.00–1763/18–00) grant to M.S. implemented as part of the TEAM-NET programme of the Foundation for Polish Science, co-financed from EU resources obtained from the European Regional Development Fund under the Smart Growth Operational Programme, a grant from the Priority Research Area “Anthropocene” under the Strategic Programme Excellence Initiative at Jagiellonian University and the Polish National Science Centre grant no. K/NCN/000514 to M.S; I.M. was supported by the National Science Centre, Poland (grant number 2023/50/E/HS6/00102) and the “Strategic research and innovation program for the development of Medical University – Plovdiv” No. BG-RRP-2.004-0007-C01.

We are very grateful to all of the children and their families for their participation in the study, to partner schools for helping us reach out to children assigned to the control group, to field psychologists for identifying children with ADHD and performing psychological testing on all children, to the study team for technical, logistic, administrative and communication efforts, and in particular to Mariola Piotrowska for invaluable organizational support throughout the project.

