## Supplementary materials for "Early-life exposure to air pollution alters resting-state functional connectivity patterns in late childhood"

**rs-fMRI data acquisition**

Resting state fMRI data were acquired using a multiband-accelerated EPI sequence (multiband factor = 6) with the following parameters: 60 axial slices, matrix size = 90 × 90, voxel size = 2.4 × 2.4 × 2.4 mm³, field of view = 216 × 216 mm² (100% phase FOV), TR = 800 ms, TE = 30 ms, and flip angle = 52°. No parallel imaging or partial Fourier was used. Two sessions of resting state fMRI data were recorded, each lasting 6.08 minutes. Prior to data acquisition, participants were familiarised with the scanner and trained to remain still using a mock scanner during a 10-minute session.

**Preprocessing**

Standard neuroimaging data preprocessing was performed using the standardized fMRIPrep (87) and CONN pipelines (88). Then, additional preprocessing steps were performed to comply with the requirements for CPM. Two-step motion correction was performed as follows. First, individual data volumes were scrubbed if their framewise displacement was larger than 0.5 mm according to the Jenkinson method (89), and participants with less than the volume equivalent of 5 minutes of data post-scrubbing were removed from further analysis. Second, the mean post-scrubbing Jenkinson framewise displacement threshold for each participant was set to 0.2 mm.

**Connectome-based predictive modeling**

In connectome-based predictive modeling (CPM), a first set of linear regressions is used to identify functional connections that are consistently associated with the variable of interest (e.g. IQ, or air pollution exposure) across participants. A summary value of those connections is calculated for each participant and used in cross-validation to predict the value of the target variable based on the connectivity. Then, a correlation between actual and predicted values of the target variables allows evaluation of model performance (89).

Model performance was quantified as the Pearson correlation between observed and predicted exposure values across participants. To assess statistical significance, this correlation was compared against a null distribution obtained through 1,000 permutation tests, in which exposure values were randomly shuffled across participants before rerunning the entire CPM procedure. The empirical p-value thus represents the proportion of permutations yielding equal or higher correlation under the null hypothesis of no association. Finally, as false positives are limited by the CPM cross-validation structure, no additional correction for multiple comparisons was applied to account for the use of multiple pollutants and exposure windows (31).

**Impact of air pollution exposure on functional network alteration**

We investigated the impact of air pollution exposure during those developmental windows for which the CPM model showed significant predictive performance, that is, where predicted and observed exposure values were significantly correlated across participants. Using predefined functional networks (default mode, cingulo-opercular, dorsal attention, ventral attention, salience, frontoparietal, medial parietal, parieto-occipital, sensorimotor, visual, auditory, and an unassigned group “none”, based on the Gordon parcellation (52), as well as a subcortical (53) and a cerebellar (54) network), we calculated the percentage of functional connections in each network that were predictive of air pollution exposure in the given period. Specifically, we used the set of functional connections retained in the final CPM masks, i.e. connections that showed significant correlations with exposure in the training folds, and counted how many of these connected ROI belong to each functional network. These percentages were computed both overall and separately for positive and negative rsFC values.

**Impact of air pollution exposure on functional network segregation and integration**

In an exploratory analysis, we assessed whether the observed impact of air pollution exposure on rsFC disrupts typical rsFC segregation and integration patterns. For each network, we computed the arithmetic mean of within-network connectivity as the average of all connectivity values between regions within the network, and the mean of between-network connectivity as the average of all connectivity values between regions in the network and all other networks. Within-network and between-network connectivity were calculated separately for positive and negative rsFC values. This decision was motivated by the logic of the segregation formula:

Segregation = Within-network / (Within-network + Between-network)

The separation allowed avoiding situations in which both numerator and denominator include negative values and thus break down the interpretation of the segregation measure. Consequently, all resulting segregation metrics were bounded between 0 and 1. A value closer to 1 indicated more within-network connectivity relative to between-network, suggesting strong segregation. Conversely, a value closer to 0 indicated more between-network connectivity relative to within-network, suggesting more integration. Division-by-zero cases (e.g., when both within- and between-network values for a given subject and network were 0) were handled by assigning NaN to the resulting segregation value and excluding it prior to model fitting.

To assess associations between brain network segregation metrics and air pollution exposure, linear regression models were fit for each pairwise combination of a given segregation metric as the predictor, and air pollution exposure as the dependent variable. The same covariates were included in the linear regression model as in the CPM. All models were fitted using Python 3.11 and the statsmodels package (ols function from statsmodels.formula.api). For each regression model, we extracted the unstandardized regression coefficient (β), standard error (SE), 95% confidence interval (CI), and p-value. We also calculated R-squared (R²) and adjusted R-squared (adj. R²) to evaluate model fit. Statistical significance was defined at p < 0.05. The results of this exploratory analysis are reported in Supplementary Table 1.

Supplementary Table 1. Association between functional network segregation at age 10-13 and exposure to PM_10_ at age 2-4.

| Network | β | r^2^ | SE | ci_lower | ci_upper | p_uncorr | p_FDR |
| --- | --- | --- | --- | --- | --- | --- | --- |
| DMN | -73 | 0.04 | 20 | -114 | -33 | 0.000 | 0.000 |
| VAN | -51 | 0.03 | 20 | -91 | -11 | 0.012 | 0.072 |
| CON | -54 | 0.02 | 25 | -103 | -6 | 0.028 | 0.112 |
| MP | -13 | 0.02 | 7 | -26 | 0 | 0.053 | 0.134 |
| VIS | -26 | 0.02 | 13 | -52 | 1 | 0.056 | 0.134 |
| PO | -11 | 0.01 | 9 | -28 | 6 | 0.204 | 0.245 |
| SUB | 12 | 0.01 | 12 | -13 | 37 | 0.335 | 0.335 |
| AUD | 10 | 0.01 | 17 | -24 | 44 | 0.557 | 0.557 |
| FP | -10 | 0.01 | 18 | -46 | 26 | 0.580 | 0.580 |
| DAN | -8 | 0.01 | 19 | -46 | 30 | 0.671 | 0.671 |
| SM | 6 | 0.01 | 22 | -37 | 49 | 0.792 | 0.792 |
| SAL | 0 | 0.01 | 6 | -12 | 12 | 0.973 | 0.973 |

β – regression coefficient, r^2^ – proportion of variance explained by the model, SE – standard error of β, ci_lower / ci_upper – 95% confidence interval for β, p_uncorr – uncorrected p-value, p_FDR – p-value corrected for false discovery rate, DMN – default mode, CON – cingulo-opercular, DAN – dorsal attention, VAN – ventral attention, SAL – salience, FP – fronto-parietal, MP – medio-parietal, PO – parieto-occipital, SM – sensorimotor, VIS – visual, AUD – auditory, SUB – subcortical. PM_10_ – particulate matter with a diameter of 10 μm or less.

Supplementary Table 2. Results of sensitivity analyses for significantly predicted PM_10_ exposure at ages 0-2 and 2-4.

| Predictive performance of CPM for PM_10_ exposure | birth-2 years | | 2-4 years | |
| --- | --- | --- | --- | --- |
|  | R | p_1000_ | R | p_1000_ |
| Main CPM model | 0.101 | 0.043 | 0.148 | 0.005 |
| SES | 0.101 | 0.039 | 0.150 | 0.007 |
| Household type | 0.103 | 0.032 | 0.149 | 0.005 |
| Early-life smoking exposure | 0.102 | 0.030 | 0.148 | 0.004 |
| Maternal alcohol use during pregnancy | 0.102 | 0.028 | 0.148 | 0.003 |

SES – socioeconomic status, assessed as the minimum education level of the two parents, Household type – living in a single-parent or two-parent household, Early-life smoking exposure – in child’s home from birth until age of 5, PM_10_ – particulate matter with a diameter of 10 μm or less, CPM – connectome-based predictive modeling, R – the correlation coefficient between the actual air pollutant exposure for the given period and the air pollutant exposure predicted by the CPM, p_1000_ – the empirical p-value for the null hypothesis R = 0 and is based on 1000 iterations of permutation testing.
